# Plasma Lipid Alterations Track Multidimensional Psychosis Severity Across Diagnostic Boundaries

**DOI:** 10.64898/2026.02.24.26346956

**Authors:** Anoja Thanabalasingam, Ariane Wiegand, Jente Meijer, Dominic B. Dwyer, Eva C. Schulte, the PsyCourse Study

## Abstract

**Background:** Lipidomic alterations have been reported across schizophrenia (SCZ) and bipolar disorder (BD), but findings are heterogeneous and often overlap across diagnoses, limiting diagnostic specificity. Associations between lipid profiles and illness severity have also been inconsistent when assessed using single symptom scales, raising the possibility that unidimensional measures fail to capture biologically relevant variation. Whether plasma lipidomic alterations relate to multidimensional psychosis severity, and how they relate to polygenic liability, remains unclear.

**Methods:** We examined associations among psychiatric and cognitive polygenic risk scores (PRS), plasma lipidomics (361 species across 16 classes), and a machine-learning-derived severe psychosis probability score in a transdiagnostic cohort of individuals with SCZ or BD (PRS n=1,320; lipid subset n=428). Regression and lipid class enrichment analyses tested severity associations. Mediation and canonical correlation analyses assessed integrated genetic–lipid–severity relationships.

**Results:** SCZ-PRS (positive), BD-PRS (negative), and educational attainment PRS (negative) showed modest associations (β = |0.02|) with severe psychosis probability. Lipid class enrichment analysis identified nine classes associated with severity, including increased sphingolipids (dSM, dCer), phosphatidylcholines (PC), triacylglycerides (TAG), and phosphatidylethanolamine plasmalogens (PE-P), alongside decreased phosphatidylcholine plasmalogens (PC-P). Most lipid class associations were robust to adjustment for diagnosis and medication. No significant mediation or shared multivariate genetic–lipid structure was observed.

**Conclusions:** Plasma lipidomic variation tracks multidimensional psychosis severity across diagnostic boundaries. These findings suggest that lipidomic alterations may reflect transdiagnostic biological processes linked to illness burden that are not fully captured by categorical diagnoses, single symptom scales, or common-variant polygenic risk.

## Introduction

Severe mental health disorders like schizophrenia (SCZ) or bipolar disorder (BD) can present with overlapping symptoms like hallucinations, delusions and disorganized thinking (1). This overlap in psychotic symptomatology motivates efforts to identify biomarkers that reflect the underlying pathophysiology of psychotic disorders that can improve diagnostic accuracy, mechanistic understanding and, ultimately, treatment stratification (2–4).

Both SCZ and BD are highly heritable with estimates between 0.6-0.8 (2), with evidence for a partially shared liability across the broader spectrum of psychotic disorders (5). Polygenic risk scores (PRS) have emerged as powerful tools for capturing this genetic liability across psychiatric disorders, offering a continuous measure of inherited risk based on genome-wide association study (GWAS) findings (6). SCZ and BD not only predict their respective diagnoses but also demonstrate transdiagnostic associations with SCZ PRS predicting psychotic features across diagnostic boundaries and BD PRS showing associations with both manic and psychotic symptomatology (7). Beyond these primary psychiatric PRS, broader batteries incorporating genetic risk for cognitive traits, personality dimensions, and other psychiatric conditions have revealed shared genetic architecture underlying diverse clinical presentations (8,9). However, while PRS show robust associations with categorical diagnoses in large population samples, they typically demonstrate much weaker effects when predicting symptom severity, functional outcomes, or treatment response with already-diagnosed individuals (10,11). This discrepancy suggests that additional biological processes may influence and track illness severity and clinical features of psychotic symptoms independently of, or in interaction with, genetic liability.

Lipid metabolism has emerged as a candidate biological pathway linking genetic vulnerability to clinical presentation in psychotic disorders given the overlap between susceptibility loci for neuropsychiatric disorders and genes linked to lipid level regulation. Lipids are key structural components of cellular membranes and signaling molecules (12), serve as energy substrates (13), comprise a substantial proportion of the brain tissue (14–16) and are implicated in synaptic plasticity, neuroinflammation, oxidative stress, and neurodevelopmental processes (14,17–20). Accumulating evidence implicated lipid alterations in psychotic disorders, including reduced plasmalogen levels in the prefrontal cortex of individuals with SCZ and altered phospholipid profiles in first-episode patients. Peripheral lipidomic studies have reported dysregulation of specific lipid classes – including sphingolipids, glycerophospholipids, and cholesterol esters.

However, findings are sometimes heterogeneous in terms of directionality of lipid level changes and often show substantial overlap across disorders, limiting diagnostic discriminatory power (18,22). This overlap suggests that lipidomic variation may index transdiagnostic pathophysiological processes rather than diagnosis-specific mechanisms. Moreover, associations between lipid profiles and illness severity have been inconsistent when assessed using single symptom scales (e.g., PANSS) (22,27,28). This may reflect the inadequacy of unidimensional measures to capture the multifaceted nature of psychotic illness severity, rather than a true absence of lipid-severity relationships.

A complementary approach to transdiagnostic research involves identifying clinically homogenous subgroups as biological exemplars against which broader phenotypic variation can be assessed. Machine learning methods applied to multidimensional data can reveal naturalistic patient groupings that may map more closely onto biological processes than traditional diagnostic categories (29,30). For instance, we previously integrated a large set of clinical variables identifying five stable psychosis subtypes in the PsyCourse study, including a severe psychosis subtype characterized by predominantly schizophrenia diagnoses, lower educational achievement, male sex, low verbal intelligence, high psychotic symptom load (but not depressive or manic symptoms), and low Global Assessment of Functioning scores (31). This subtype’s clinical homogeneity and symptom severity make it an optimal target for biomarker discovery, as extreme phenotypes typically demonstrate larger biological effect sizes than broader diagnostic groups. Critically, the machine learning model generates a continuous probability score reflecting each individual’s simillarity to this severe subtype, creating a multidimensional severity measure.

Despite growing evidence that lipid metabolism is altered in psychotic disorders and that psychiatric PRS capture transdiagnostic genetic liability, no study has examined whether peripheral lipidomic alterations are associated with multidimensional psychosis severity or whether such alterations mediate the relationship between genetic risk and clinical presentation. Understanding whether lipid changes reflect heritable biological vulnerability (trait markers) or illness state processes has important implications. If lipid alterations mediate genetic risk, they represent potential mechanisms linking genomic variation to clinical outcomes and may suggest targetable pathways for intervention. Conversely, if lipid changes track with illness severity independently of genetic liability, they may reflect state-dependent processes - such as neuroinflammation, oxidative stress, or metabolic dysfunction - that emerge during acute illness or accumulate over illness course.

To address these questions, we investigated the relationships among plasma lipidomic profiles, polygenic risk scores, and multidimensional psychosis severity in a transdiagnostic cohort of the PsyCourse Study. We hypothesized that: (1) specific lipid classes would show enrichment in individuals with higher severe psychosis subtype probability; (2) psychiatric PRS would demonstrate associations with this severity measure; and (3) genetic liability and lipid alterations would show integrated relationships with illness severity. We tested this using two complementary approaches: mediation analysis examines whether specific lipid alterations transmit genetic risk to clinical outcomes (a sequential pathway model), while canonical correlation analysis assesses whether coordinated genomic-lipidomic variation - independent of individual associations - relates to severe psychotic presentation (a shared variation model). We tested these hypotheses using 13 psychiatric and cognitive PRS and machine-learning-derived severe psychosis probability in 1320 individuals with schizophrenia spectrum or bipolar disorders. Within this cohort, 428 participants had comprehensive plasma lipidomics (361 species, 16 classes), enabling integrated genetic-lipidomic analyses including mediation and canonical correlation approaches.

## Methods

### Participants

The PsyCourse Study (https://www.psycourse.de)(version 6.0) is a longitudinal German-Austrian deep-phenotype study including participants with severe mental disorders and healthy controls. A subset of 1320 participants who were diagnosed according to DSM-IV with SCZ, brief psychotic disorder, schizoaffective disorder, or BD were included in this analysis. Genomic data was available for all n=1320 included participants, with additional lipidomic data available for subset of n=428 hereof. Genetic analysis was performed with the full sample (n=1320) and lipidomic and integrated analysis on the subset of n=428. Written informed consent was obtained from each participant. The study was approved by the University Hospital Munich’s ethics committee (Project number: 17-13) and is compliant with the Declaration of Helsinki.

### Severe Psychosis Subtype Probability

We used a machine learning-derived severity measure based on a previously identified severe psychosis subtype from the PsyCourse cohort, version 4. This subtype was characterized by elevated positive symptoms (hallucinations, delusions), cognitive impairment, and functional disability, identified through clustering of multidimensional clinical assessments. Following the identification of five distinct clinical subtypes through clustering, we trained a support vector machine (SVM) classifier using a sparse set of clinical variables selected for optimal discrimination between subtypes.

For the present analysis, we required continuous severity scores applicable to all participants rather than categorical cluster assignments in the PsyCourse cohort version 6.0. We therefore replicated the SVM classifier on the current sample (n=1,320) using the same sparse variable set. Probability scores for severe psychosis subtype membership were obtained using Platt scaling (scikit-learn CalibratedClassifierCV with 5-fold internal cross-validation), providing a continuous measure ranging from 0 (low probability) to 1 (high probability). The probability assigned to the severe psychosis subtype (cluster 5) served as our primary outcome measure for all analyses. The replicated SVM successfully reproduced the severe psychosis subtype classification with comparable performance metrics (see Supplementary Methods and Table S1 for validation).

### Genotyping and Polygenic Risk Scores

Participants were genotyped using the Illumina Infinium Global Screening Array-24 Kit (GSA Array, version 1 and 3; Illumina, San Diego, CA). Details about the pre-processing pipeline including quality control and imputation (HRC [Version r 1.1. 2016] reference panel) has been reported in (32). Briefly, we employed a hypothesis-driven PRS battery encompassing psychiatric liability (SCZ (33), BD (34), major depressive disorder (MDD) (35)), cognitive function (Alzheimer’s disease (36), educational attainment (37)), neurodevelopmental disorders (ASD (38), ADHD (39)), which show substantial genetic overlap with psychotic disorders and elevated comorbidity in severe psychosis cohorts (8), and traits relevant to psychosis pathophysiology (Big Five personality traits (40) and sleep duration (41)(42,43). Polygenic risk scores (PRS) were derived from the summary statistics of recent large-scale genome-wide association study (GWAS). For each GWAS, variants were retained if they (1) had an imputation quality score ≥ 0.6 (when available), (2) had minor allele frequency ≥ 0.01 in the 1000 Genomes Phase 3 European reference panel, and (3) were present in both the GWAS summary statistics and the imputed PsyCourse genotype data. We applied PRS-CS (44) using the 1000 Genomes European LD reference to obtain posterior single-nucleotide polymorphism (SNP) effect sizes. PRS-CS was run using default parameters and a grid of pre-specified global shrinkage (φ) values (1×10⁻⁶, 1×10⁻⁵, 1×10⁻⁴, 1×10⁻³, 1×10⁻², 1×10⁻¹) to capture varying degrees of polygenicity. For protective variants, alleles were re-oriented so that weights were consistently aligned to the trait-/risk-increasing allele. PRS were then computed from these weights and the imputed target-cohort genotypes using PLINK 1.9 and R (v.4.0). To construct lipid-metabolism-enriched psychiatric PRS, we used the REACTOME_METABOLISM_OF_LIPIDS gene set from the Molecular Signatures Database (MSigDB v2023.2; (45)), which contains 700 genes involved in lipid biosynthesis, transport, storage, and catabolism. Lipid-related genes were mapped to genomic coordinates using GENCODE (GRCh37) and for each psychiatric GWAS, we restricted SNPs to those located within the ± 50kb region of these lipid genes. This window size captures promoters and proximal enhancers while maintaining gene-specificity, consistent with widely-used gene-set analysis approaches (46). PRS-CS were always run on the full set of HapMap3 SNPs for each GWAS to estimate genome-wide posterior effect sizes. Lipid-specific PRS were then obtained by restricting the PLINK scoring step to the subset of SNPs in proximity to lipid genes. All PRS were standardized before inclusion in downstream models.

### Lipidomics

Non-fasting plasma samples were collected between 2013 and 2016 and analyzed between 2018 and 2020. Lipid quantification was performed using liquid chromatography coupled with untargeted mass spectrometry (LC-MS) as previously described (22). Briefly, samples were analyzed on a Waters Acquity UPLC system coupled to a Q Exactive orbitrap mass spectrometer (Thermo Fisher Scientific) with heated electrospray ionization. Lipids were separated using reverse-phase chromatography (ACQUITY UPLC BEH C8 column, 2.1 × 100 mm, 1.7 μm) and mass spectra were acquired in both positive and negative ionization modes. Peak detection and integration were performed using XCMS software with the “centWave” method (47). Of 1361 detected lipid features, 394 were annotated to lipid species using an in-house spectral library, spanning 16 lipid classes: triacylglyceride (TAG), acylcarnitine (CAR), phosphatidylcholine (PC), phosphatidylcholine plasmalogen (PC-P), ceramide (Cer), phosphatidylethanolamine (PE), phosphatidylethanolamine plasmalogen (PE-P), fatty acid (FA), sphingomyelin (SM), plasmanylphosphatidylcholine (PC-O), cholesteryl ester (CE), diacylglycerol (DAG), lysophosphatidylcholine (LPC), lysophosphatidylcholine plasmalogen (LPC-P), lysoplasmanylphosphatidylcholine (LPC-O), and lysophosphatidylethanolamine (LPE).

For the present analysis, we applied sequential quality control exclusions: (i) 31 lipid species with extreme distributional properties (skewness| ≤ 1, |kurtosis| ≤ 5) (ii) 30 lipid species known to be affected by fasting status (primarily fatty acids) (39). After accounting for overlap between these categories, 361 lipid species across 16 lipid classes were retained. Individual outlier values (z-score > 3 within each lipid species) were set to missing prior to analysis. Lipid intensities were log2-transformed and z-score standardized across individuals. Lipid class scores were computed as the mean of z-scored species within each class.

### Statistical Analysis

#### Regression Analysis

To test association between severe psychosis subgroup probability and PRS, we performed an ordinary least squares linear regression analysis using robust standard errors (HC3) with the covariates age, sex, and the first five ancestry principal components, correcting for multiple testing using the Benjamini-Hochberg (BH) correction. Lipid species association analysis was performed analogously to the PRS association analysis, sex, body mass index (BMI), duration of illness and smoking status as covariates. Both PRS and lipid regression analysis was assessed for sensitivity by adding the covariates diagnosis and number of medications.

##### Lipid Class Enrichment Analysis

We tested whether lipid classes showed coordinated associations with severe psychosis using a gene set enrichment analysis (GSEA) approach adapted for lipidomics. Lipid species were ranked by their signed regression coefficients (β) from the univariate models (capturing both effect magnitude and direction). An enrichment score was computed for each lipid class containing ≥5 species. Statistical significance was assessed using 5,000 permutations in which lipid coefficients were randomly shuffled while preserving class membership structure. We report normalized enrichment scores (NES; normalized by the mean and standard deviation (SD) of permutation scores with the same sign), empirical p-values, and BH FDR-corrected q-values across all tested lipid classes.

#### Statistical Mediation Analysis

To test whether lipid alterations statistically mediated the association between PRS and severe psychosis, we performed linear mediation analyses in the subset with available lipidomics with the continuous probability of belonging to the severe psychosis subgroup as the dependent variable. Separate mediation models were run for each PRS (e.g. PRS-SCZ (or PRS_SCZ_), PRS-BD, PRS-MDD, and their lipid-metabolism-enriched counterparts), entered as the independent variable (X) for each mediator (M), e.g. lipid class that was significantly enriched in previous lipid class enrichment analysis. PRS were residualized for age, sex, and the first five ancestry principal components. Lipid class values were residualized for age, sex, BMI, duration of illness and smoking status. Covariate-adjusted residuals were used in all subsequent analyses. Mediation models were estimated with ordinary least squares using pingouin v.0.5.5, yielding estimates of the direct effect (X→Y), indirect effect (X→M→Y) and total effect (sum of direct and indirect effects). Confidence intervals and p-values for indirect effects were obtained via non-parametric bootstrap with 5000 resamples.

#### Canonical Correlation Analysis

Canonical correlation analysis (CCA) identifies linear combinations of variables from two domains that exhibit maximal correlation, revealing coordinated patterns of variation. We tested shared variation between PRS and lipid classes using two-block (CCA) using scikit-learn. 13 PRS and 16 lipid class features were residualized separately for covariates (PRS: age, sex, PC1-PC5; lipids: age, sex, BMI, smoking, duration of illness), median/mode-imputed for missing values, and z-standardized. We fit CCA (1 component) on the residualized blocks and assessed the first canonical correlation via permutation (10,000 label permutations of the lipid block). The resulting PRS and lipid canonical scores were then tested for association with severe psychosis probability using HC3-robust OLS, with permutation-based p-values (20,000 outcome permutations). Canonical loadings were reported to characterize contributing features.

Significance level was alpha = 0.05. All analysis was performed with Python 3.12 with the following packages: Numpy (v.2.3.0), pandas (v.2.3.0), scikit-learn (v.1.7.0), statsmodels (v.0.14.4),SciPy (v.1.15.2), pingouin (v.0.5.5), matplotlib (v.3.10.3) and seaborn (v.0.13.2). Analysis code can be accessed under https://github.com/anthano/psycourse.

## Results

### Participant Characteristics

**Table 1.**
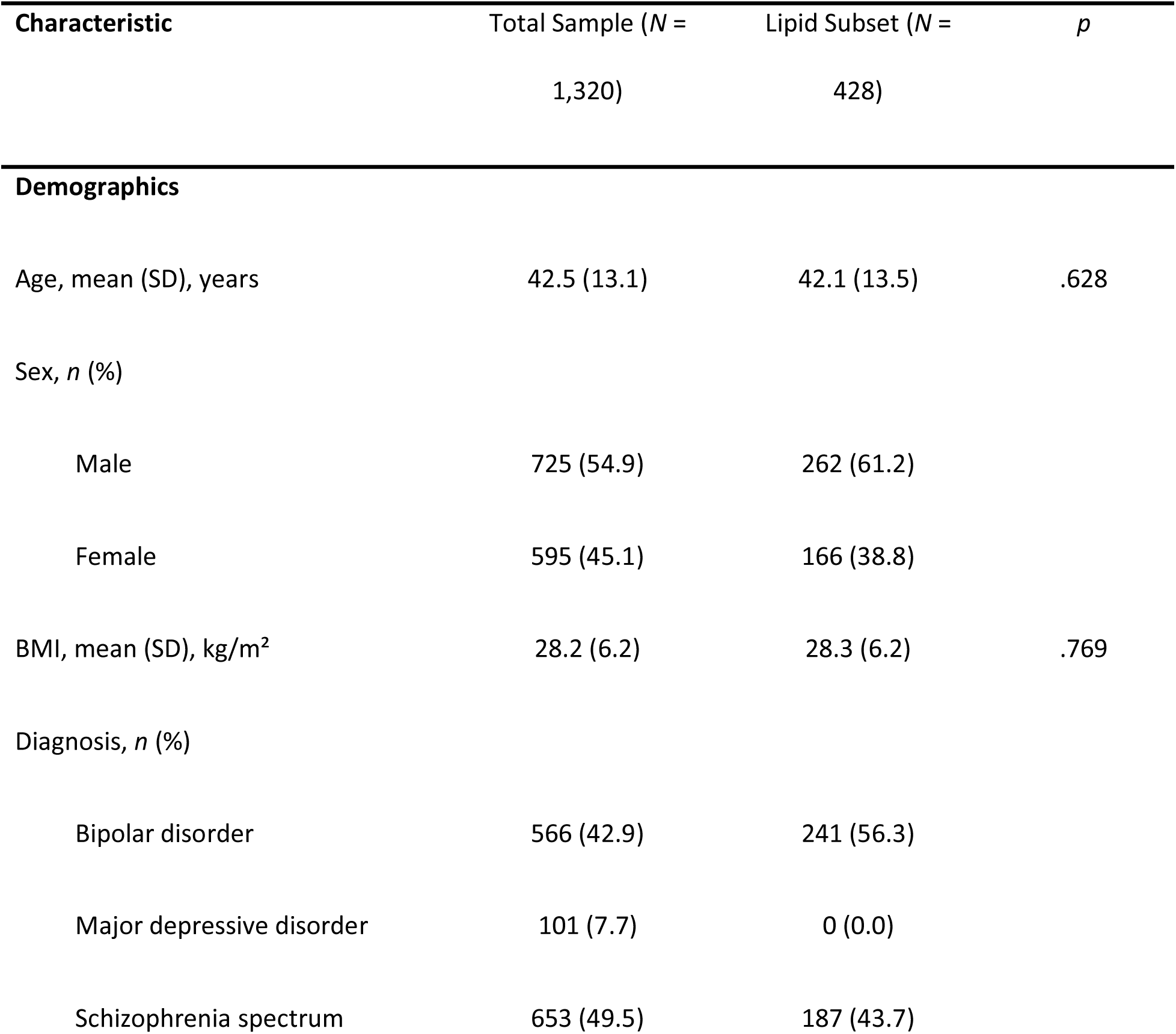
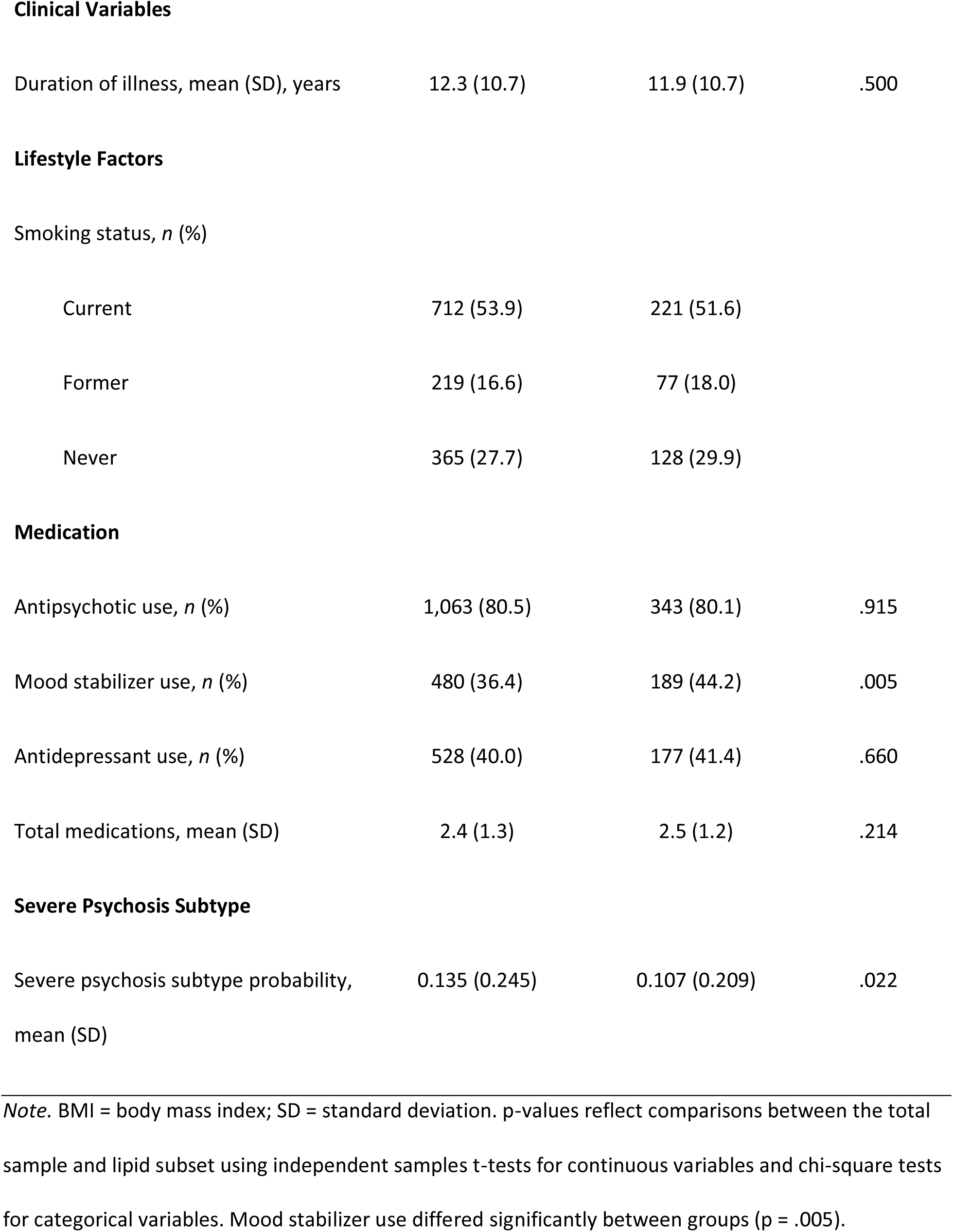
Demographic and Clinical Characteristics. Demographic, Clinical, and Medication Characteristics of the Total Sample and Lipid Subset

### PRS Associations

Three out of 13 tested PRS showed significant associations with severe psychosis subtype probability: higher SCZ-PRS (β=0.02, q=0.045), lower BD-PRS (β=-0.02, q=0.02), and lower educational attainment PRS (β=-0.02, q=0.045) (Figure 1.

**Figure 1.**
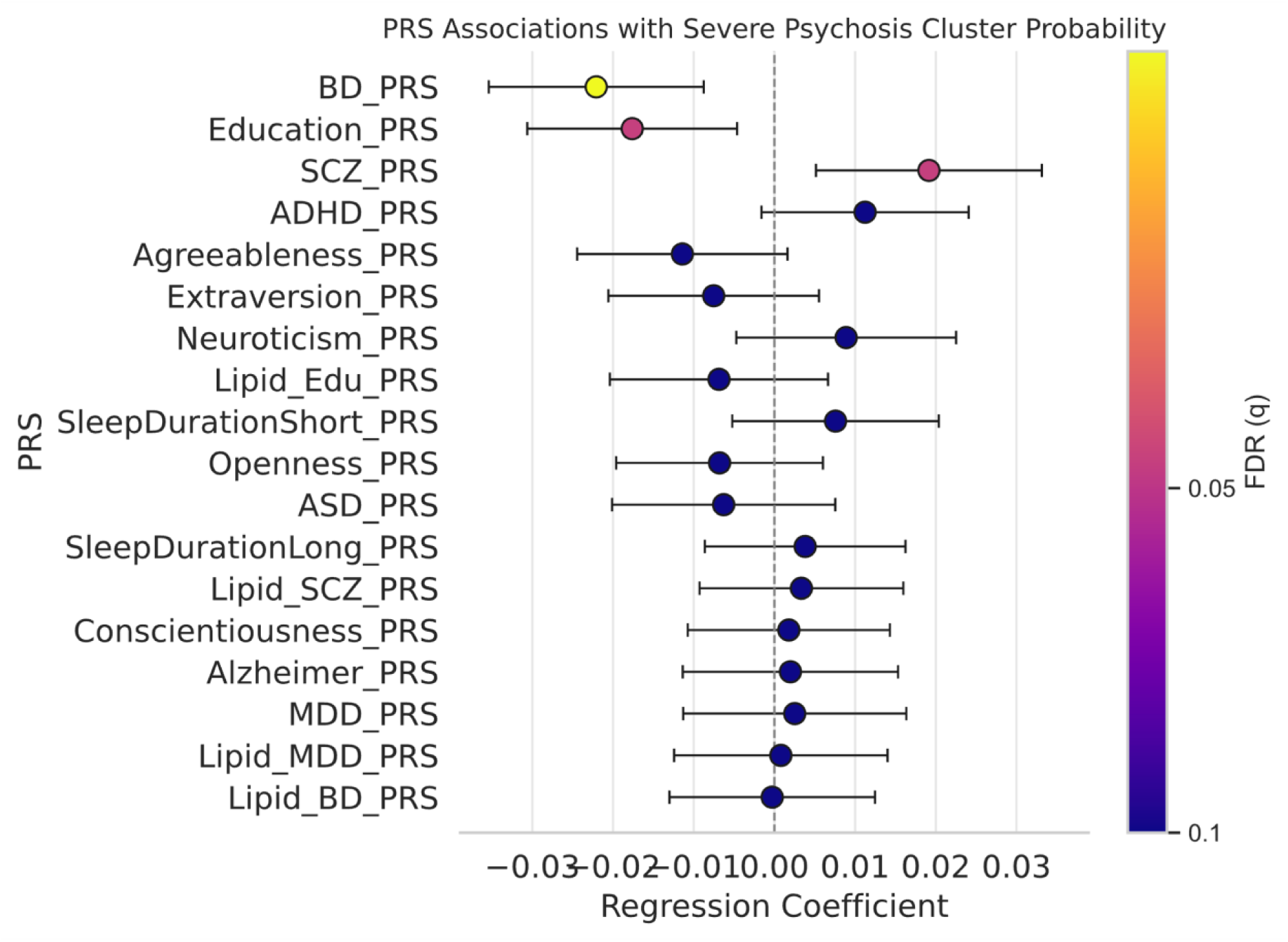
Psychiatric PRS are associated with severe psychosis subtype. OLS coefficients with 95% CIs (heteroskedasticity-robust), adjusted for covariates (age, sex, BMI, Top 5 ancestry PCs) components. Colors = −log10(FDR); colorbar ticks = FDR (q) values.

After controlling for diagnosis, all PRS associations were attenuated and no longer statistically significant, indicating that PRS differentiate diagnostic groups but provide limited additional discrimination of transdiagnostic psychosis severity in this sample (Supplemental Figure 3). Lipid-restricted psychiatric PRS (SNPs within ±50kb of lipid genes) showed no significant associations with severe subtype probability in either primary or diagnosis-adjusted models (all q>0.05; Table S4-6).

### Lipid Associations with Severe Psychosis Subtype

#### Individual Lipid Species

Before controlling for diagnosis and medication, three lipid species were significantly associated with severe psychosis subtype probability after FDR correction: PE-P 42:5 (β=0.05, q=0.002), PE-P 40:4 (β=0.03, q=0.013), and PE 40:5 (β=0.05, q=0.033)(Supplemental Figure 5). All three belong to the phosphatidylethanolamine plasmalogen (PE-P) class. After controlling for diagnosis and medication, no individual species remained significant (Supplemental Figure 6, Supplemental Figure 7).

#### Lipid Class Enrichment

In contrast to individual species, lipid class enrichment analysis revealed robust associations. In the primary model (adjusting for age, sex, BMI, smoking, duration of illness), nine lipid classes showed significant enrichment (q<0.05). The sphingolipids classes dSM (NES=1.577, q=0.002) and dCer (NES=1.285, q=0.003), as well as the Glycerophospholipid classes PC (NES=2.663, q=0.001), PE-P (NES=1.285, q=0.003), PE (NES=1.245, q=0.018), and TAG (NES=1.112, q=0.001), CE (NES=1.904, q=0.005), PC-O (NES=1.656, q=0.018) were positively enriched with severe psychosis probability. PC-P (NES=-1.383, q=0.001) was negatively associated with the severe psychosis probability (Figure 2. Most enrichments persisted after adjusting for diagnosis (8 of the 9 classes) or medication (7 of the 9 classes) in a sensitivity analysis, though PC-O reversed direction with both adjustments, suggesting diagnosis- or treatment-related confounding for this class (Figure 2. When adjusting for both diagnosis and medication, we observed positive enrichment in PC, TAG, dSM, PC-O and PE-P, while we observed negative enrichment in PC-P, PC-O and LPE (Supplemental Figure 9; Supplemental Table 10-13).

**Figure 2.**
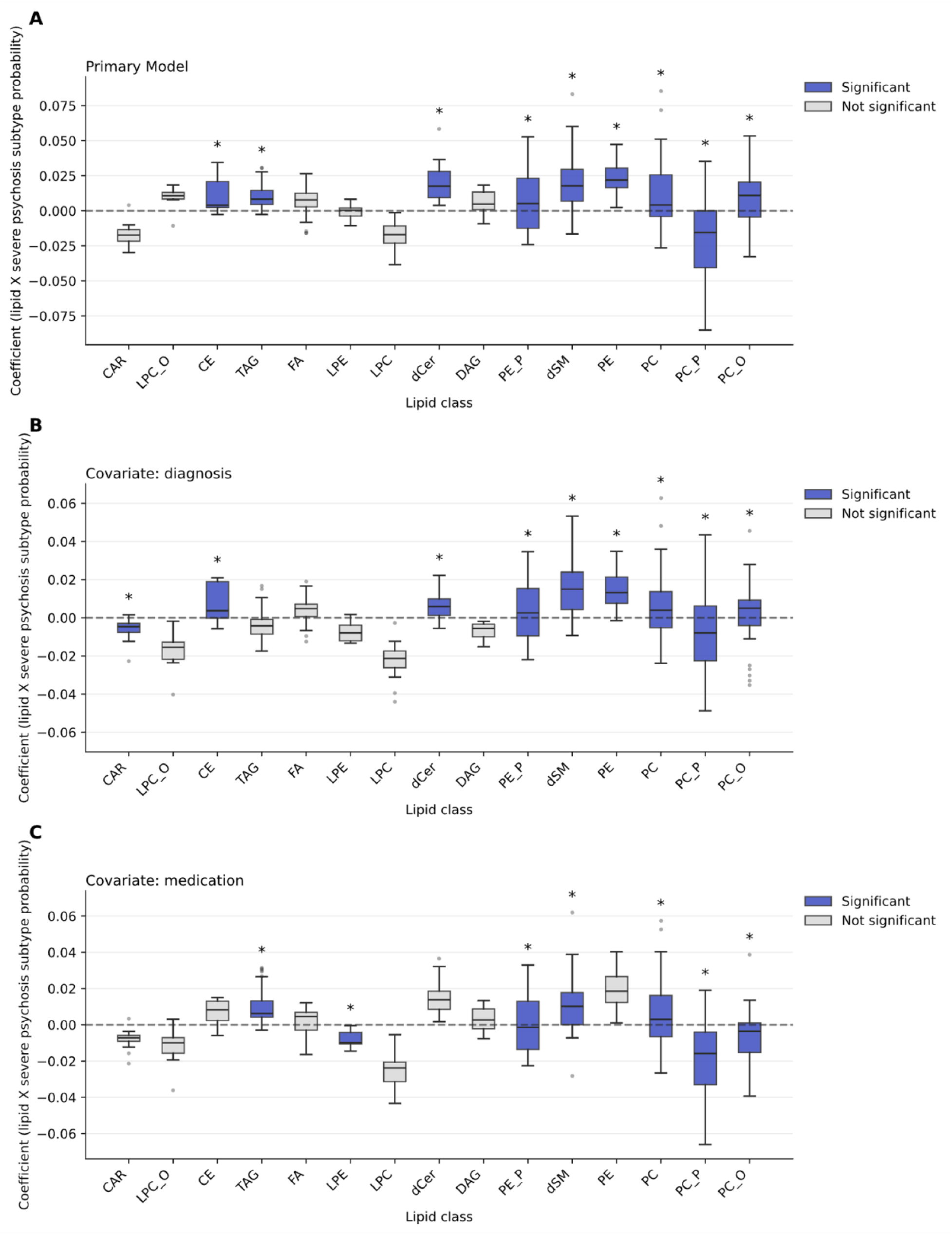
Lipid-class based enrichment analysis for severe psychosis subtype probability. Boxplots of ordinary least squares regression coefficients (β) for individual lipid species, grouped by lipid class. Each box represents the distribution of species-level coefficients within a class. Blue boxes indicate lipid classes with FDR-significant enrichment (q < 0.05) from permutation-based gene set enrichment analysis (5,000 permutations); grey boxes indicate non-significant classes. Asterisks denote significantly enriched classes. The dashed horizontal line indicates β = 0. Outliers shown as individual points. **(A)** Primary model, adjusted for age, sex, BMI, smoking status, and duration of illness; **(B**) additionally adjusted for diagnosis; **(C)** additionally adjusted for number of medications.

### Mediation of Genetic Liability Through Lipid Alterations

We tested whether enriched lipid classes mediated associations between psychiatric PRS and severe psychosis subtype using bootstrap-based mediation analysis. We examined indirect effects for three psychiatric PRS (SCZ-PRS, BD-PRS, MDD-PRS) through nine enriched lipid classes from the initial lipid class enrichment analysis.

None of the 27 tested mediation pathways showed significant indirect effects (all bootstrap 95% CIs included zero; (Supplemental Figure 10)). The largest indirect effect was for SCZ-PRS → dSM → severity (β=0.003, 95% CI: -0.001 to 0.009, p=0.18).

### Shared Genetic-Lipidomic Variation

In a post hoc CCA between residualized PRS and lipid-class profiles (1 component), the first canonical correlation was modest (r_c = 0.33) and not significant by permutation testing (p_perm = 0.604; 10,000 permutations). Neither the PRS nor lipid canonical score was associated with severe psychosis probability (HC3 OLS; β = -0.0003 p_perm = 0.978 and β = - 0.0068 p_perm = 0.741, respectively). Loadings were diffuse; the largest absolute contributors were Openness, Short Sleep Duration, BD-PRS on the PRS side and DAG, CE, PE-P, CAR, PE on the lipid side (|loading| ≤ 0.4), and are reported descriptively. This pattern suggests limited shared covariation between common-variant genetic liability and peripheral lipidomic variation in this sample (Figure 3).

**Figure 3.**
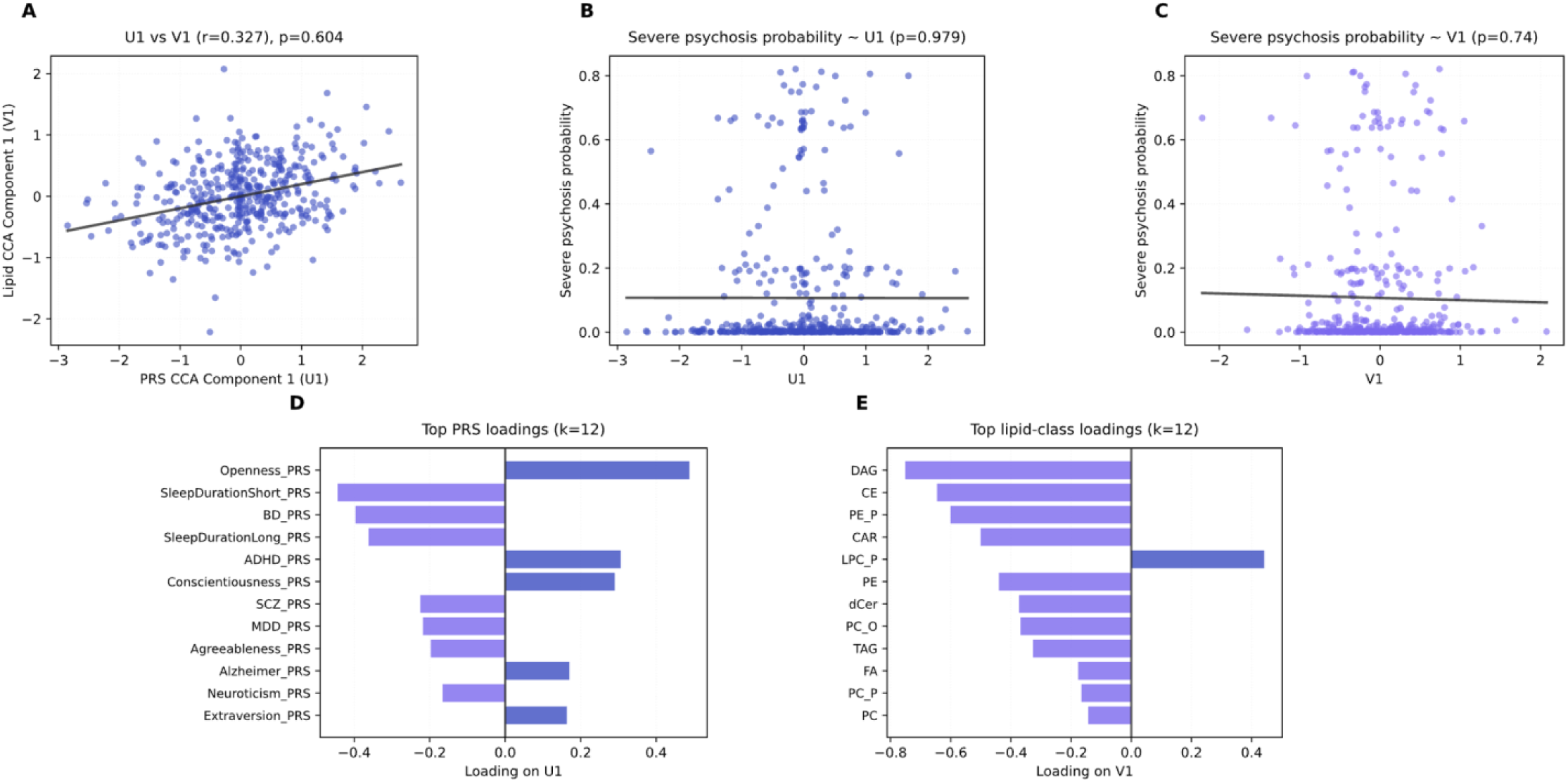
Canonical correlation analysis of polygenic risk scores and lipid classes. **(A)** Relationship between the first PRS canonical component (U1) and the first lipid canonical component (V1); canonical correlation was modest (r=0.33, p=0.604, permutation test, 10,000 permutations). **(B, C)** Association of PRS (U1) and lipid (V1) canonical scores with severe psychosis subtype probability (β=−0.0003, p=0.979 and β=−0.007, p=0.740, respectively). **(D, E)** Canonical loadings showing contributions of individual PRS variants **(D)** and lipid classes **(E)** to their respective canonical components; all loadings |r|≤0.40, indicating diffuse rather than class-specific contributions. Together, the null associations and modest canonical correlation suggest limited shared genetic-lipidomic covariation with illness severity.

## Discussion

In this study, we molecularly characterized a severe psychosis subgroup using plasma lipidomics and PRS, in a sample spanning SCZ and BD diagnoses.

Three PRS showed significant associations with severe psychosis probability: higher SCZ-PRS (β=0.02, q=0.045), lower BD-PRS (β=-0.02, q=0.02), and lower EA-PRS (β=-0.02, q=0.045). The positive SCZ-PRS association indicates that genetic liability for schizophrenia predicts transdiagnostic psychotic severity. The negative EA-PRS association aligns with cognitive impairment as a core feature of severe psychosis and with known inverse genetic correlations between cognitive ability and schizophrenia (48). The negative BD-PRS association suggests that bipolar genetic liability - which captures risk for manic and affective features - may be orthogonal or even protective against severe psychotic presentations. This pattern is consistent with evidence that psychotic features in BD are driven by SCZ-PRS rather than BD-PRS (7) and that BD with psychosis shows genetic overlap with schizophrenia (49). The combination of positive SCZ-PRS and negative BD-PRS association in our severe subtype suggests genetic architecture more aligned with schizophrenia spectrum biology than with affective components of bipolar disorder, regardless of DSM-5 diagnosis. The modest effect sizes are consistent with prior work showing that PRS generally explain less variance in continuous clinical severity or treatment outcomes than in case-control disease status (50). This likely reflects both the inherent complexity of severity phenotypes and the fact that available PRS were derived from GWAS of categorical diagnoses rather than continuous psychotic symptom severity, for which large-scale GWAS do not yet exist.

We identified enrichment of glycerophospholipid (PC, PE-P, PE) and sphingolipid classes (dSM, dCer) associated with severe subtype probability. Within glycerophospholipids, enrichment was characterized by increases in plasmalogen-containing phosphatidylethanolamines (PE-P) alongside decreases in plasmalogen-containing phosphatidylcholines (PC-P), suggesting headgroup-specific ether-lipid remodeling. At the species level, two plasmalogens (PE-P 42:5, PE-P 40:4) showed the strongest positive associations with psychosis. Notably, ether-linked PC-O enrichment attenuated and reversed direction after adjustment for diagnosis or medication, suggesting that PC-O changes may primarily reflect diagnosis-specific or treatment-related variation, highlighting the importance of covariate-adjusted models.

Plasmalogens are glycerophospholipids characterized by a vinyl-ether bond at sn-1 and are abundant in myelin and synaptic membranes, where they influence membrane dynamics, vesicle fusion, and serve as oxidative buffers (51,52). Plasmalogen dysregulation has been documented across peripheral and brain tissues in psychotic disorders, with functional consequences including cognitive deficits and neuroinflammation (53–55). Peripheral studies report 23-45% reductions in plasma choline and ethanolamine plasmalogens in SCZ (56), with similar decreases observed in drug-naive first-episode patients (57), indicating that these alterations are not solely attributable to antipsychotic exposure. Brain tissue studies add further complexity to this picture. In prefrontal cortex gray matter, a comprehensive lipidomic analysis identified widespread alterations in glycerophospholipid metabolism in schizophrenia, including PE and PC classes (58). Studies of prefrontal white matter report both increases and decreases in specific phospholipid species: Ghosh et al. (2017) found lower PC alongside elevated PE22:0 and PE24:1 in bipolar disorder, with decreased n-6 polyunsaturated fatty acids (PC20:4n6, PE22:5n6, PC22:5n6) in both schizophrenia and bipolar disorder. These variable findings across studies likely reflect methodological differences in lipid detection and quantification (targeted vs. untargeted lipidomics), tissue compartment differences (gray vs. white matter), and the specific lipid classes examined.

Our headgroup-specific pattern (PE-P increases, PC-P decreases) aligns with evidence for differential phospholipid regulation in severe psychosis and may reflect selective modeling of ether lipids under oxidative and metabolic stress. PE-P upregulation could represent a compensatory antioxidant response, whereas reduced PC-P may reflect preferential consumption, impaired synthesis, or greater vulnerability to oxidative damage given differences in fatty acid composition and membrane distribution. Alternatively, differential regulation of plasmalogen-synthesizing or remodeling enzymes across headgroup classes may contribute to this imbalance. Notably, our observation of increased peripheral PE-P contrasts with reports of reduced brain plasmalogens in schizophrenia, underscoring that plasma lipid profiles may reflect systemic metabolic or inflammatory processes rather than direct CNS concentrations.

We observed enrichment of sphingolipid classes (dSM, dCer) associated with severe psychosis probability. Sphingolipids comprise structurally diverse lipids built on a sphingoid base, serving both as structural membrane components (sphingomyelin, cerebrosides) and bioactive signaling molecules (ceramide, sphingosine, sphingosine-1-phosphate (S1P)) that regulate cell fate, inflammation, and myelination. Central to sphingolipid biology is the “sphingolipid rheostat,” whereby ceramide and S1P exert opposing effects on cell survival: ceramide promotes growth arrest and apoptosis, while S1P supports proliferation and counters ceramide-mediated cell death. Inhibition of sphingosine kinase shifts the balance toward ceramide accumulation and triggers apoptosis, highlighting how the relationship between these metabolites regulates cell fate. In postmortem brain tissue of SCZ patients, Esaki et al. (2020) reported lower S1P levels in corpus callosum alongside higher gene expression of S1P-degrading enzymes and S1P receptors, suggesting a compensatory response to reduced S1P availability (59). This metabolic shift was linked to impaired oligodendrocyte differentiation and myelin formation, providing a possible mechanistic explanation for white matter abnormalities documented in SCZ. Translational studies combining animal models and human data have shown that changes in sphingolipid-metabolizing enzymes (acid sphingomyelinase, ceramidases, sphingomyelin synthase) in prefrontal and striatal regions track with psychosis-like behavior and its reversal with haloperidol, implicating sphingolipid enzyme activity in both pathogenesis and antipsychotic response.

Plasma studies in SCZ and BD repeatedly report elevated ceramides in untreated or baseline states (22,60). Brunkhorst-Kanaan et al. (2019) found elevated levels of ceramides and their hexosyl-metabolites in plasma samples from SCZ, BD and MDD patients (24). In another late-onset SCZ study, lysosphingolipids (e.g., HexSph, LysoSM) were elevated alongside decreased acid sphingomyelinase activity (61). In contrast, sphingomyelin findings were more variable and often treatment-dependent (62,63). Sphingolipid dysregulation is also well documented in inflammatory disorders and cancer, where the ceramide/S1P axis governs stress and immune signaling (64–66). This transdiagnostic involvement raises the possibility of therapeutic repurposing, as S1P receptor modulators (e.g., fingolimod) and functional inhibitors of acid sphingomyelinase are already clinically available (67).

In our study, ceramide enrichment remained significant after adjusting for diagnosis but was attenuated and no longer significant after adjustment for medication, whereas sphingomyelin enrichment remained robust to both adjustments. Prior work suggests susceptibility of ceramide to medication: risperidone treatment has been associated with decreases in plasma-C1P and certain sphingomyelins, particularly in poor responders (63), antidepressants have been associated with increased levels of plasma-ceramides (24), while other studies report elevated ceramide levels to be independent of antipsychotic treatment in prefrontal cortex white matter and red blood cells (68). The sphingomyelin signal’s robustness to medication adjustment suggests it may more closely reflect severity-related biology.

Interpretation of peripheral class-level changes remains challenging, as cellular outcomes depend on the balance of multiple species rather than absolute levels, as well as enzyme/receptor activity (69). The mechanistic basis for sphingolipid dysregulation in severe psychosis and its relationship to central pathophysiology remains an important question requiring studies that integrate peripheral lipidomics with neuroimaging, CSF sampling, and assessment of inflammatory and cardiovascular risk profiling.

Our findings both corroborate and extend recent case-control lipidomic studies. While we replicate core findings from a recent large-scale case-control lipidomic study from our research group (Tkachev et al., 2023) (PC-P decreases, Cer/TAG/PC increases), we identify additional enriched classes (LPE, PC-O, PE, PE-P, dSM, CE) associated with severe psychosis probability. Critically, Tkachev et al. (2023) found no lipidomic differences between high versus low PANSS severity groups, leading them to conclude lipid alterations represent trait markers. Our contrasting finding of robust lipid-severity associations using a multidimensional phenotype suggests that single symptom scales may lack sensitivity to detect severity-related lipidomic variation. The additional enriched classes in our analysis may specifically index severe, multidimensionally-defined illness burden.

To probe whether lipid alterations might represent the link between polygenic liability and severe psychosis subtype probability, we performed mediation analyses. Despite moderate SNP-based heritability of lipid classes (range between 0.0 – 0.45, with sphingomyelins and ceramides at ≈0.34–0.35) (70), suggested genetic overlap between psychiatric risk loci and lipid metabolism genes (71,72), and 14-fold enrichment of lipid genes among loci implicated in SCZ, BD, and MDD (22), we found no evidence for PRS-lipid-severity mediation. This null finding likely reflects limited statistical power (73). However, it also suggests that severity-associated lipid changes may be primarily state-dependent - driven by illness chronicity, inflammation, metabolic dysregulation, or treatment, rather than inherited liability. Larger genetic studies enriched for severe presentations are needed to test lipid-mediated pathways.

To our knowledge, this is the first application of canonical correlation analysis to integrate polygenic risk and peripheral lipidomics in severe psychosis. Although multivariate methods such as CCA and related approaches have been used to link genomic and metabolomic data in other contexts, we are not aware of prior CCA-based integration of PRS and lipidomics in psychotic disorders. While CCA and related multivariate methods (sparse CCA, multiblock PLS) have been successfully applied to link genetic, transcriptomic, and metabolomic data in cardiovascular and metabolic disorders - identifying coordinated gene-metabolite modules related to inflammation, lipoprotein metabolism, and energy homeostasis - analogous sample-level multivariate integration in psychiatry remains rare. Most published psychiatric metabolomics studies have used PLS-DA for diagnostic classification or PCA-based dimensionality reduction, while genetic-metabolite relationships are primarily examined through summary-level methods (genetic correlation, Mendelian randomization) rather than individual-level canonical variates.

We applied CCA to test whether linear combinations of PRS (n=13) and lipid classes (n=16) capture shared multivariate structure beyond univariate associations. The first canonical correlation was r = 0.33 (p = 0.604), but permutation testing (10,000 iterations) revealed this was not statistically significant, indicating the apparent correlation likely reflects sampling variability rather than true genetic-lipidomic covariation. This null finding aligns with the absence of PRS-lipid mediation and suggests that, in this cohort, common-variant polygenic liability and severity-associated lipid alterations show limited shared linear covariation - at least as captured by current PRS and peripheral lipidomics.

While the null result suggests minimal overlap in this sample, this analysis demonstrates the feasibility of applying established cardiovascular-metabolic integration methods to psychiatric cohorts with high-dimensional biological data. Future studies with larger samples, broader lipidomic coverage, and rare-variant or transcriptomic data may reveal genetic-metabolite modules associated with specific symptom dimensions or treatment response trajectories that are missed by cross-sectional peripheral lipidomics and PRS-based approaches such as those used here.

Several limitations should be noted. First, we assessed plasma lipids as a peripheral proxy for brain lipid metabolism, which may not fully reflect central nervous system processes. Second, participants were non-fasted, introducing potential confounding from recent diet and circadian variation, although this was partly mitigated by excluding lipid species known to be strongly influenced by fasting status (22). Third, although we statistically adjusted for medication effects, residual confounding from specific drugs, dosages, combinations, and overall lifestyle and environmental factors cannot be excluded. Fourth, because our polygenic scores indexed categorical diagnoses, rather than psychotic symptoms or transdiagnostic psychosis severity, they may not optimally capture genetic liability to the severe psychosis phenotype examined here, reflecting the current absence of GWAS for those dimensions. Fifth, we cannot rule out reverse causation, whereby severe psychosis and its correlates (e.g. stress, inflammatory activation, health behaviours) drive lipidomic changes, rather than lipids contributing to symptom severity. Notably, Mendelian randomization studies have begun to support at least partially causal roles for specific lipid species in BD and MDD, particularly for polyunsaturated fatty acids and phospholipids, but evidence in SCZ is more mixed and often points to modest bidirectional effects between circulating lipid metabolites and SCZ risk. To our knowledge, there are no other large datasets combining SCZ and BD, untargeted lipidomics, and detailed psychosis-severity phenotyping that would permit direct replication of our clustering and mediation analyses. Finally, although this is among the largest untargeted lipidomic studies in psychotic disorders to date, our sample size still limited power for mediation and canonical correlation analyses, and replication in larger, independent cohorts is needed.

## Conclusions

In a transdiagnostic psychosis cohort, we found that severe psychosis subtype probability - defined using machine learning integration of clinical variables - was associated with reproducible plasma lipidomic shifts, particularly in plasmalogen-containing glycerophospholipids and sphingolipids. These associations were largely robust to adjustment for diagnosis and medication, suggesting that they could reflect severity - related pathophysiology rather than diagnostic or treatment confounds. In contrast, PRS for neuropsychiatric traits provided limited discrimination beyond diagnosis, and we found no evidence that lipid alterations mediate common-variant-derived genetic liability.

These findings support the hypothesis that peripheral lipid profiles index psychosis-severity-related biology that is partly state- and environment-sensitive, rather than simply reflecting diagnosis-associated genetic liability. By leveraging a multidimensional psychosis severity phenotype, this study extends prior work using single symptom scales (22) and demonstrates that lipidomic correlates may be more robustly detected when psychosis severity is defined transdiagnostically. Future work should employ longitudinal data with large-scale lipidomics data, accounting for various confounding factors, and larger samples to clarify the temporal dynamics, prognostic utility, and mechanistic relevance of lipidomic variation in psychotic disorders.

## Supplement

**Supplemental Figure 1.**
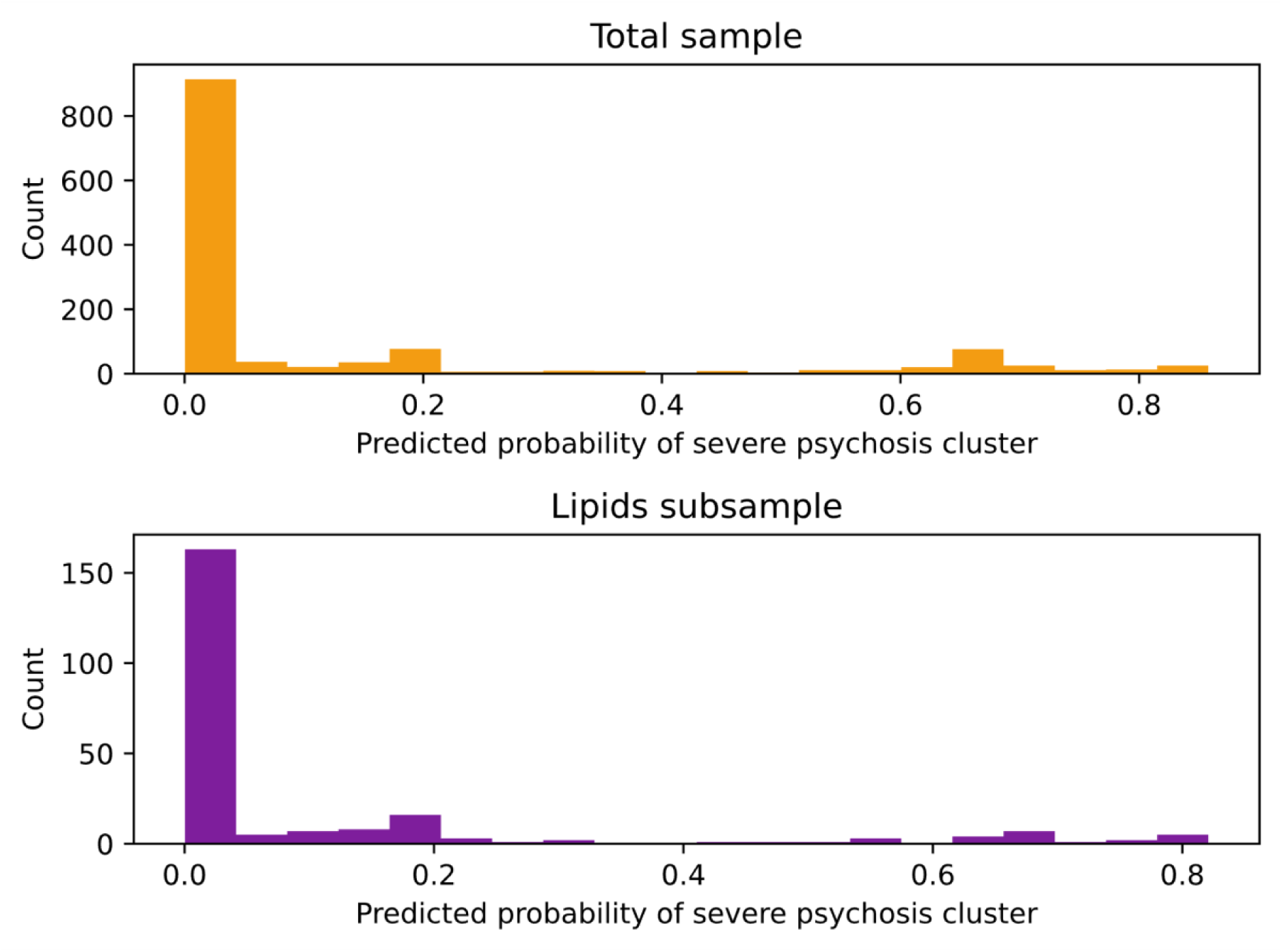
Distribution of predicted probability of severe psychosis probability in the total sample (n=1320) and the lipid subsample (n=428).

**Supplemental Figure 2.**
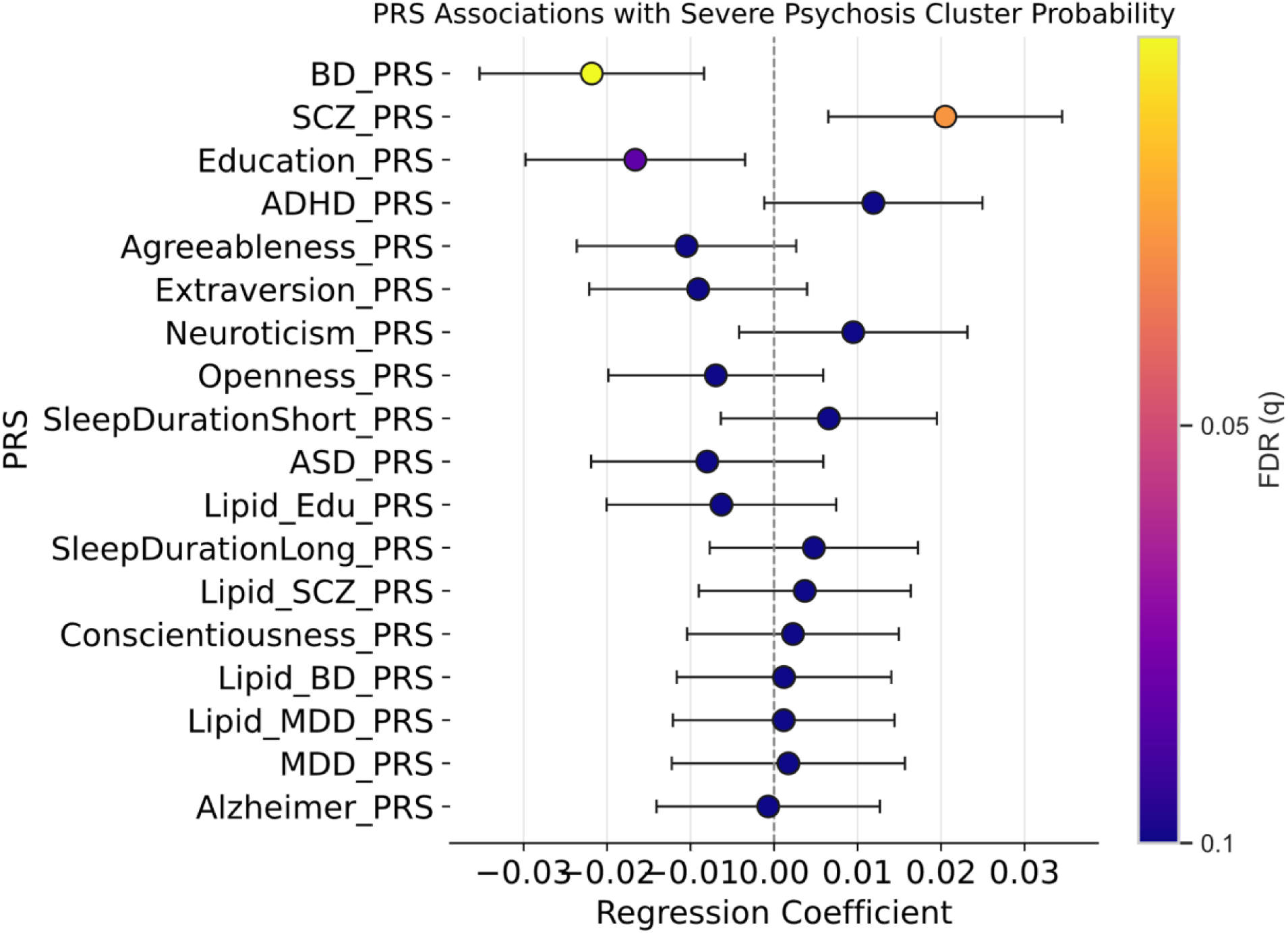
PRS Regression with BMI as added covariate. Individual PRS associations with severe psychosis probability. OLS coefficients with 95% CIs (heteroskedasticity-robust), adjusted for covariates (age, sex, BMI, Top 5 ancestry PCs) components. Colors = −log10(FDR); colorbar ticks = FDR (q) values.

**Supplemental Figure 3.**
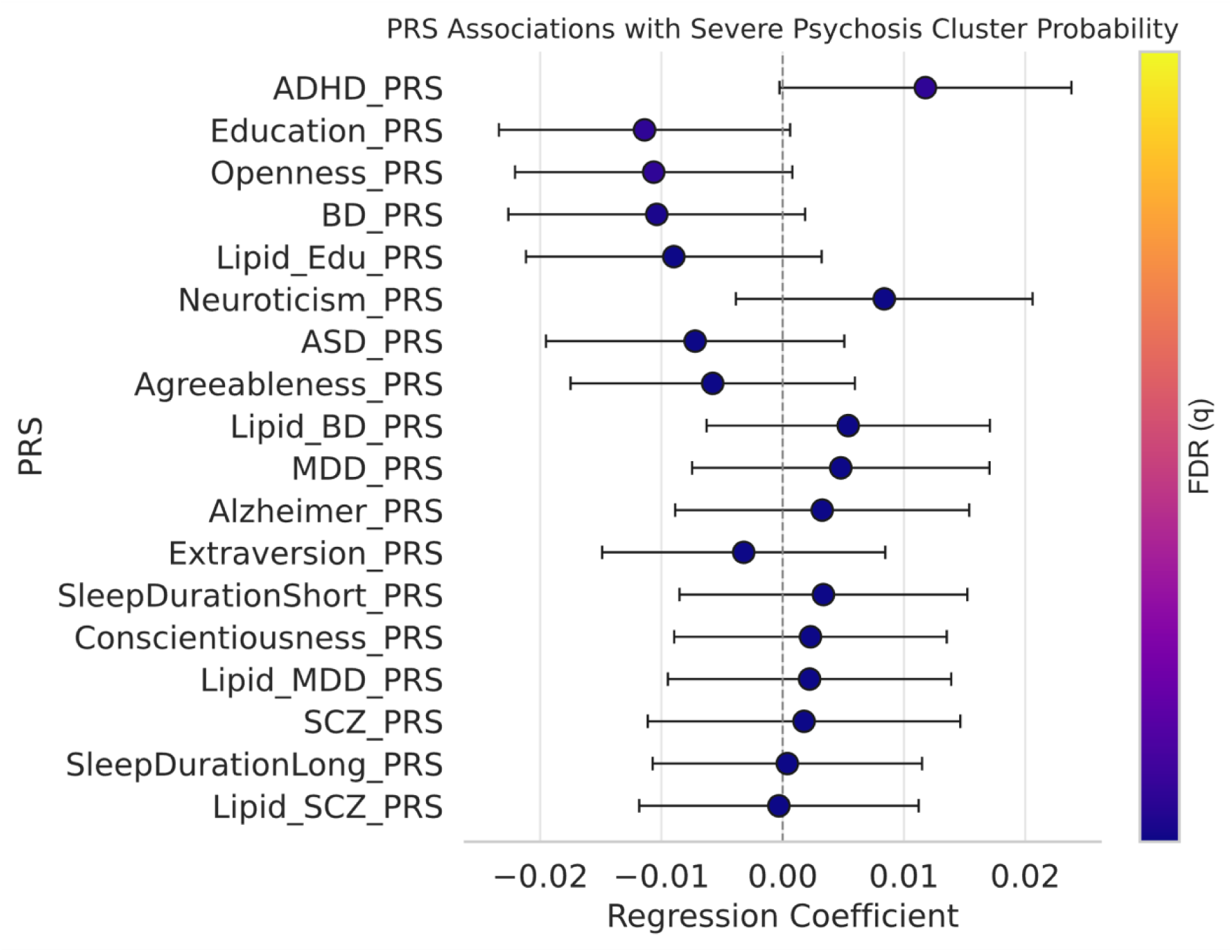
PRS Regression with diagnosis as added covariate. Individual PRS associations with severe psychosis probability. OLS coefficients with 95% CIs (heteroskedasticity-robust), adjusted for covariates (age, sex, diagnosis, Top 5 ancestry PCs) components. Colors = −log10(FDR); colorbar ticks = FDR (q) values.

**Supplemental Figure 4.**
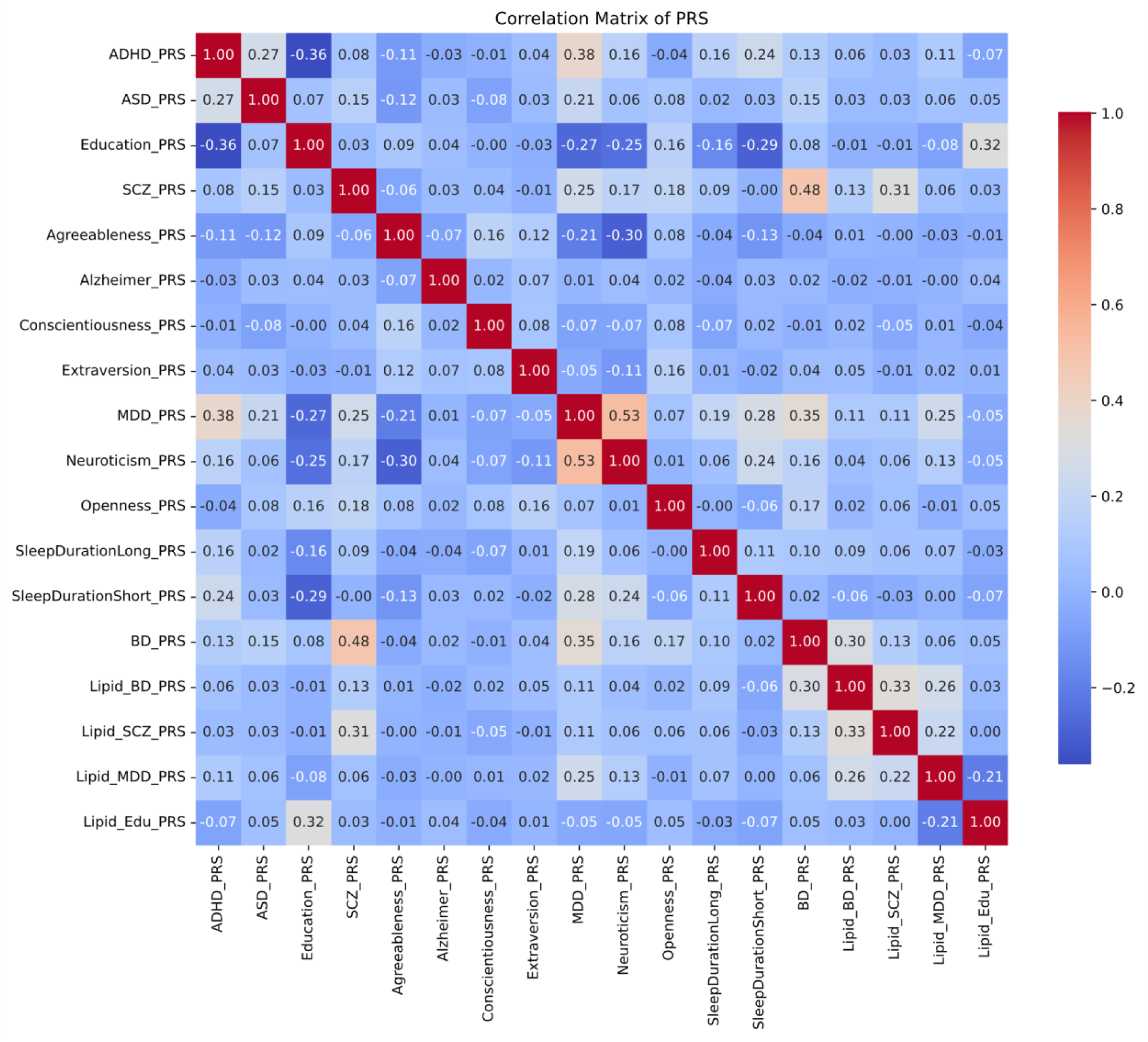
Correlation Matrix of PRS.

**Supplemental Figure 5.**
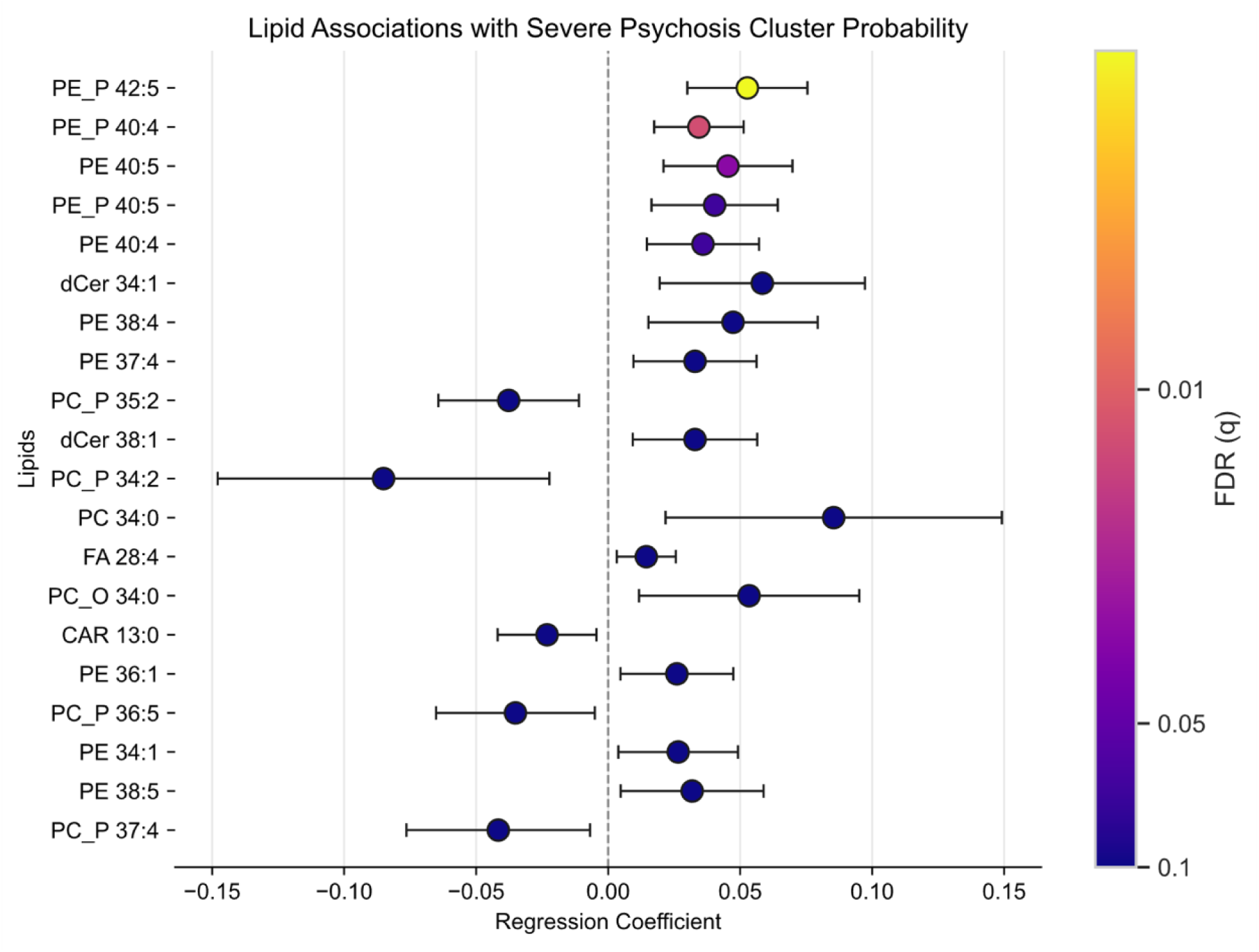
Lipid species are associated with severe psychosis subtype probability. OLS coefficients with 95% CIs (heteroskedasticity-robust), adjusted for covariates (age, sex, BMI, duration of illness and smoking status) components. Colors = −log10(FDR); colorbar ticks = FDR (q) values.

**Supplemental Figure 6.**
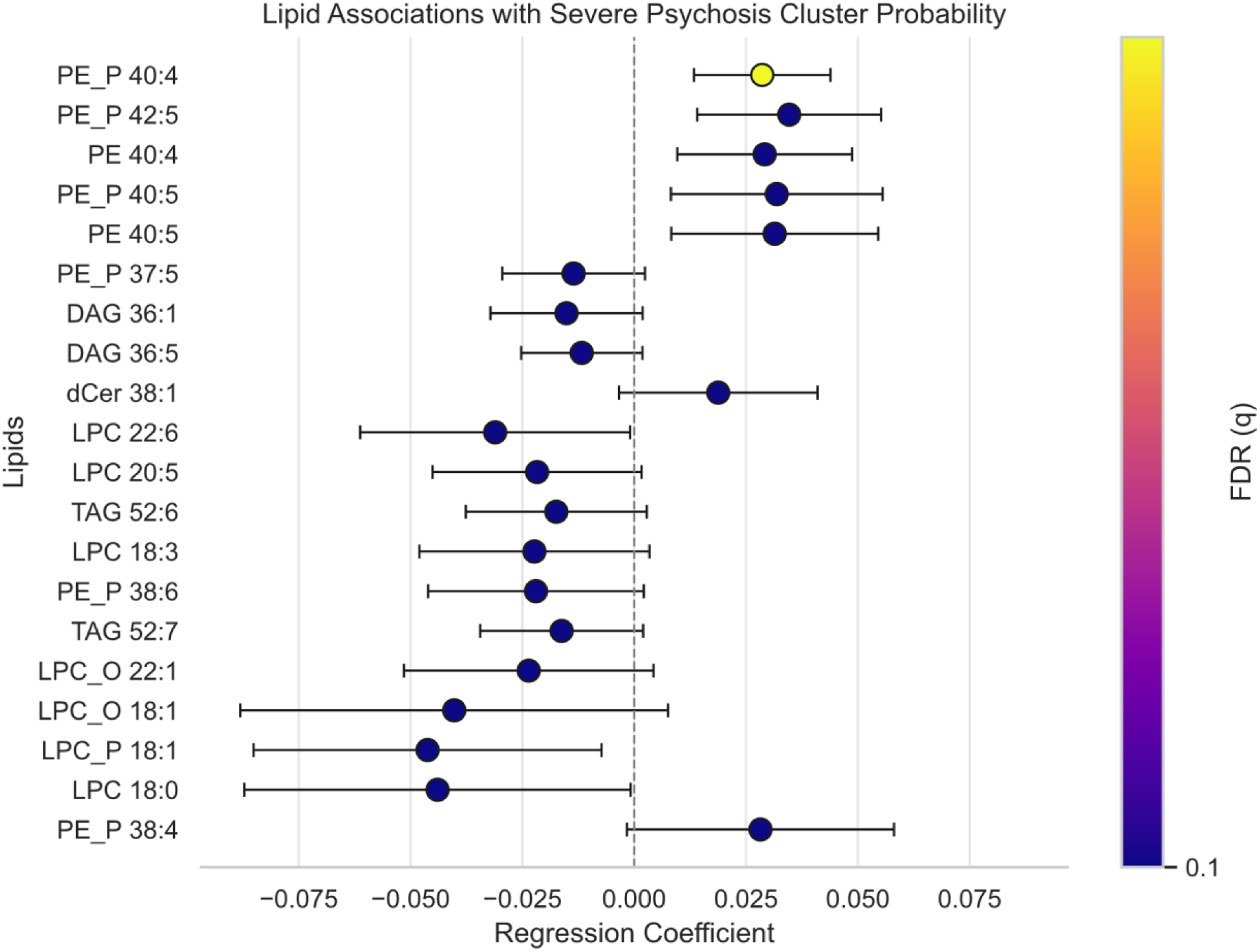
Lipid species associations with severe psychosis subtype probability after adjusting for diagnosis. OLS coefficients with 95% CIs (heteroskedasticity-robust), adjusted for covariates (age, sex, BMI, duration of illness, smoking status, and diagnosis). Colors = −log10(FDR); colorbar ticks = FDR (q) values. No lipid species survived FDR correction (all q > 0.05).

**Supplemental Figure 7.**
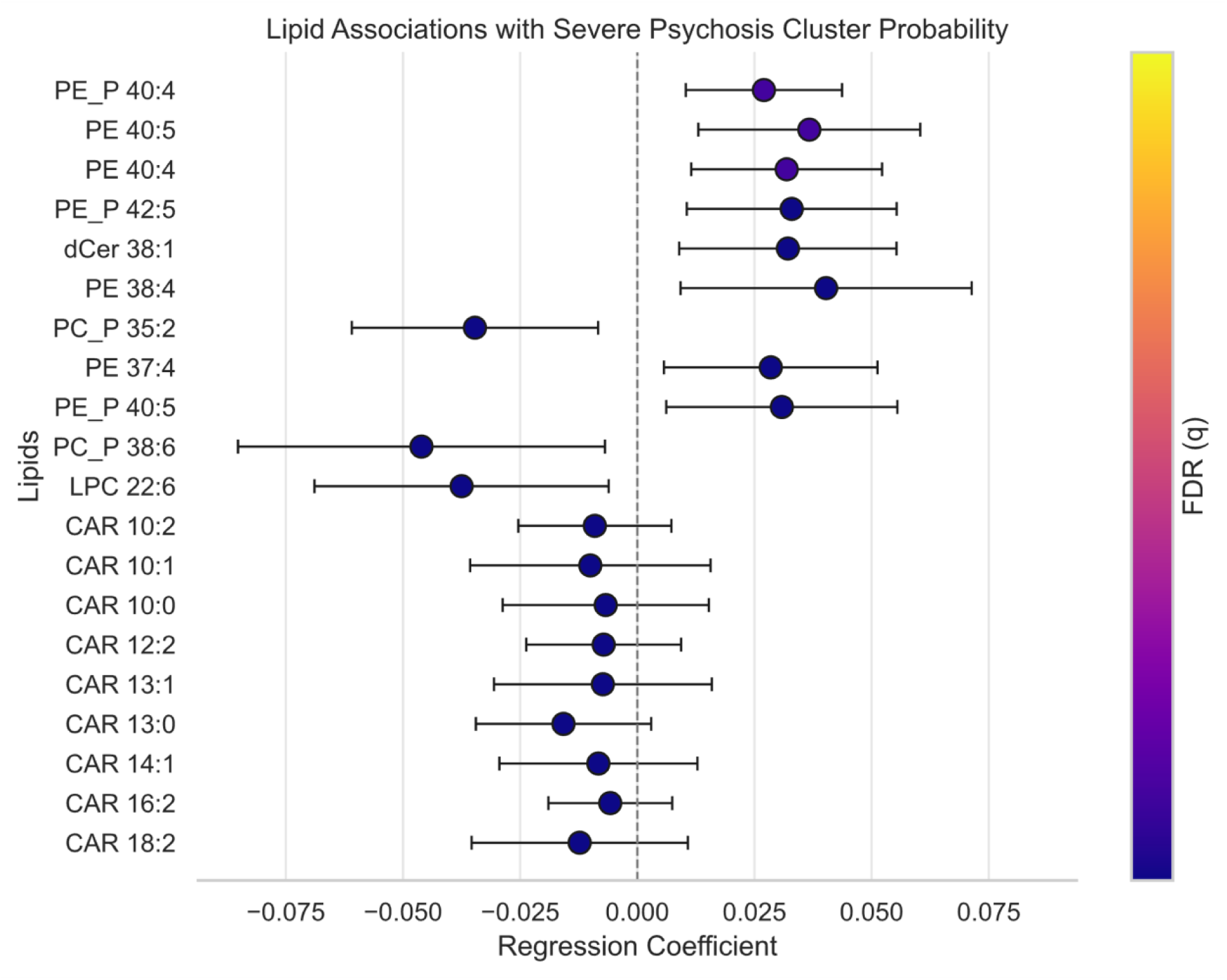
Lipid species associations with severe psychosis subtype after adjusting for medication. OLS coefficients with 95% CIs (heteroskedasticity-robust), adjusted for covariates (age, sex, BMI, duration of illness, smoking status, and medication). Colors = −log10(FDR); colorbar ticks = FDR (q) values. No lipid species survived FDR correction (all q > 0.05).

**Supplemental Figure 8.**
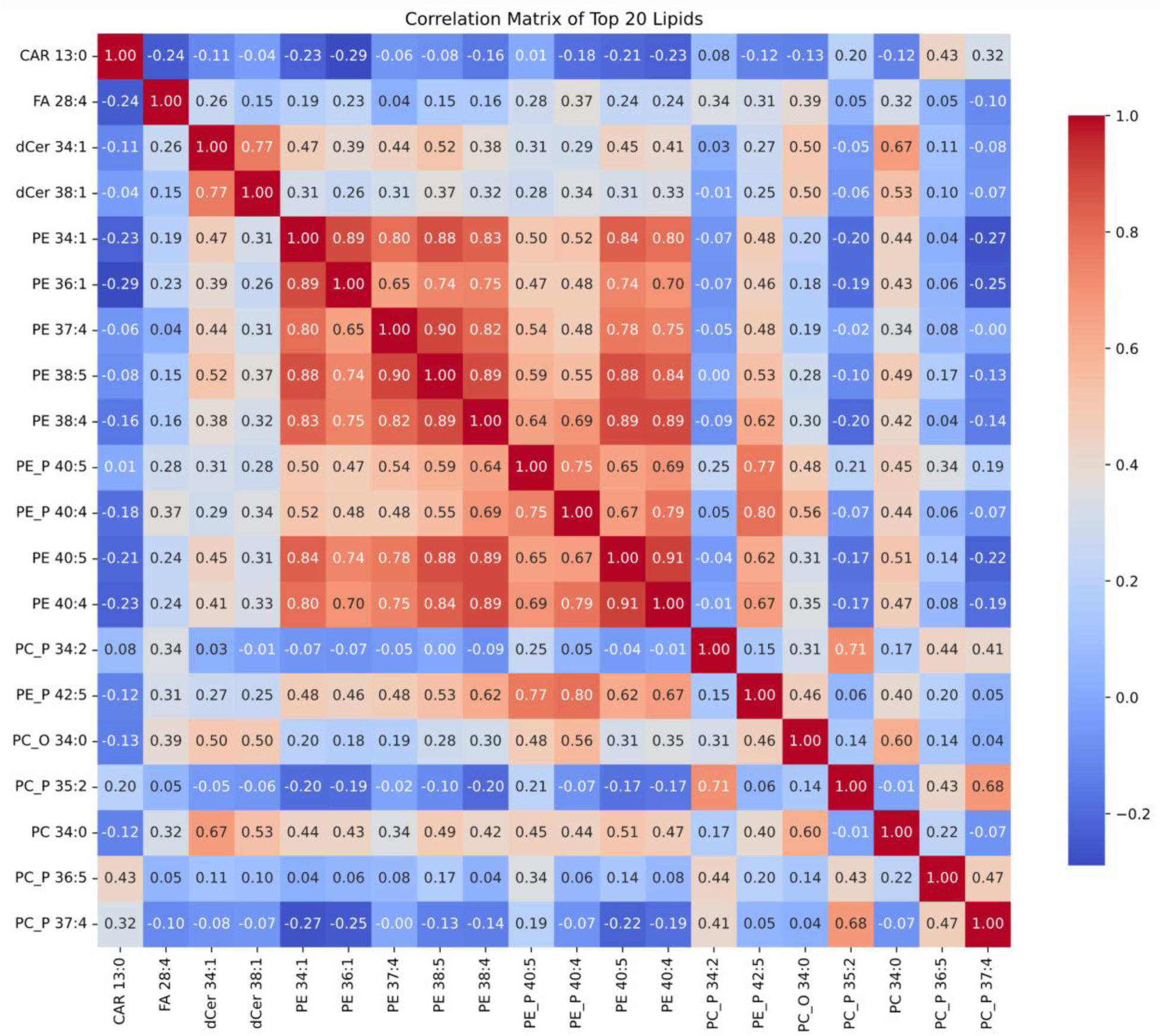
Lipid Species Correlation.

**Supplemental Figure 9.**
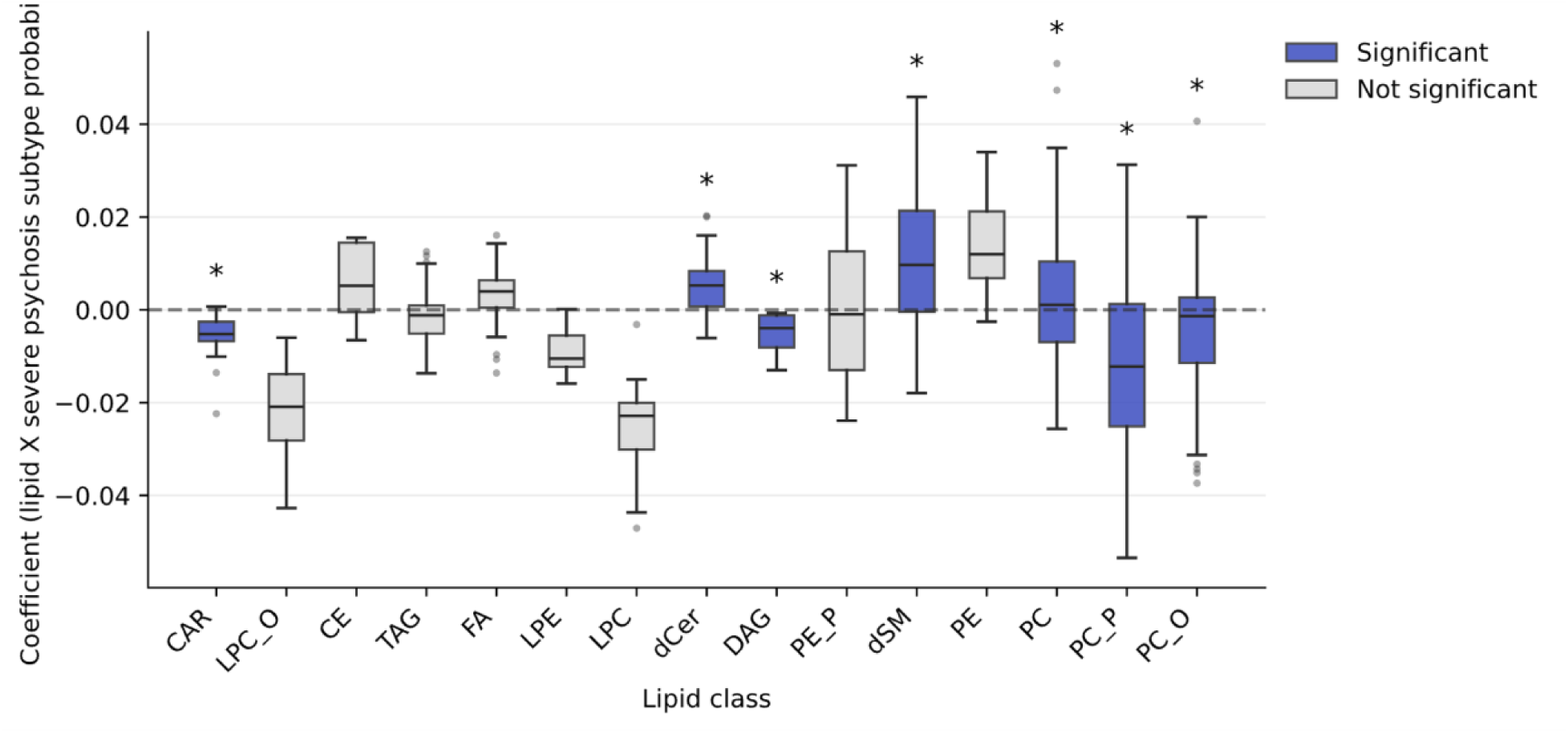
Lipid class enrichment in severe psychosis subtype with added covariates medication and diagnosis. Normalized enrichment scores (NES) from permutation-based gene set enrichment analysis. Lipid species were ranked by regression coefficients from associations with severe psychosis subtype probability (adjusted for age, sex, BMI, smoking, duration of illness). Blue bars indicate FDR-significant enrichment (q<0.05). Positive NES indicates lipids positively associated with severity; negative NES indicates inverse association. Additional covariates: number of medications and diagnosis.

**Supplemental Figure 10.**
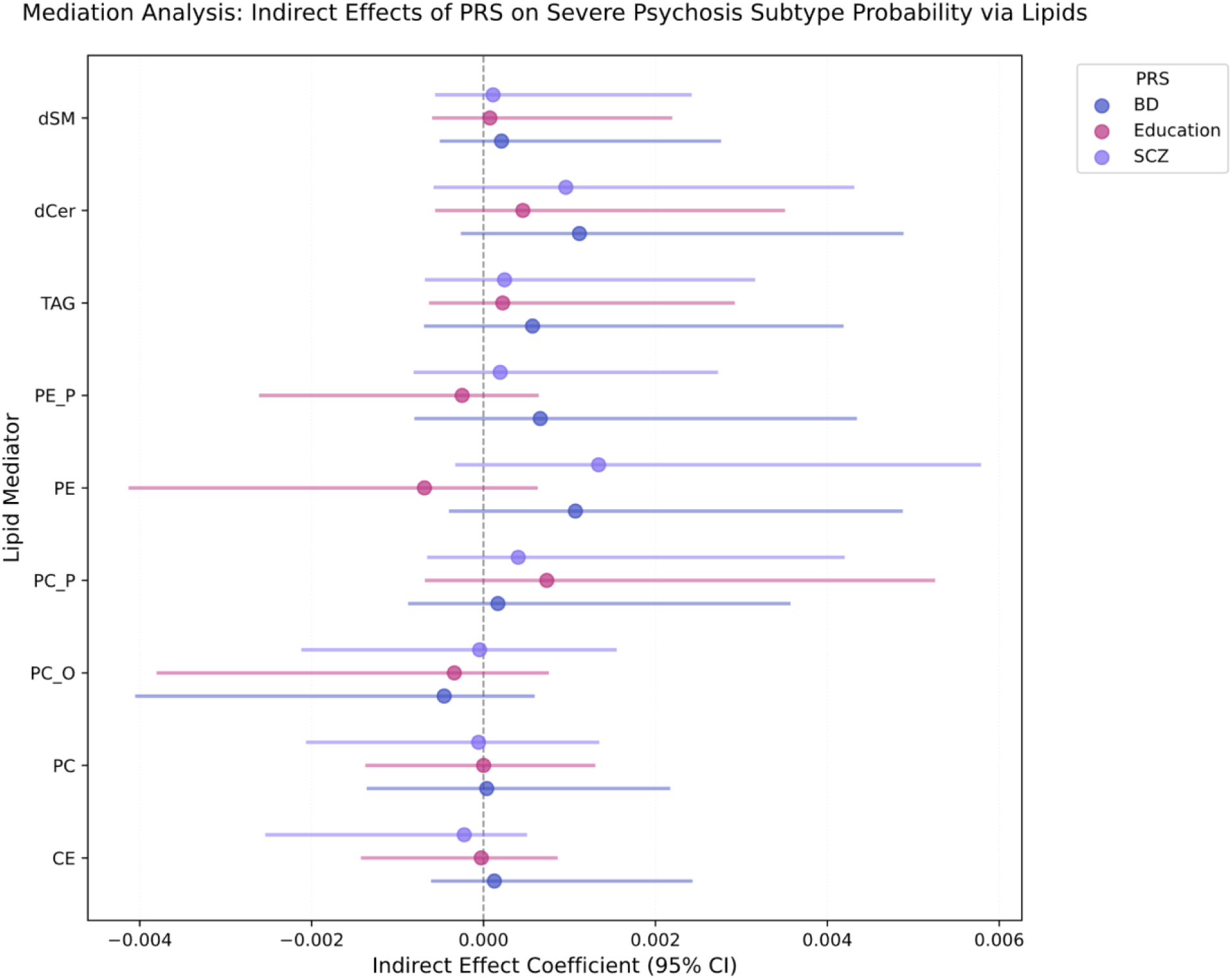
Mediation of genetic liability through lipid classes. Forest plot of indirect effects (95% CI) from bootstrap mediation analysis testing whether enriched lipid classes mediate PRS associations with severe psychosis. None of 27 pathways showed significant mediation (all CIs include zero), suggesting lipid alterations do not mediate polygenic liability.

## Data Availability

A unique feature of the PsyCourse Study is that it has been conceptualized as a continuously growing data resource available to the scientific community. Data sharing will be based on mutually agreed research proposals and within the Open Science framework of the PsyCourse Study.

https://www.psycourse.de/openscience-en.html

